# An assessment of evidence to inform best practice for the communication of acute venous thromboembolism diagnosis: a scoping review

**DOI:** 10.1101/2024.10.29.24316375

**Authors:** Samarth Mishra, Frederikus A. Klok, Grégoire Le Gal, Kerstin de Wit, Aviva Schwartz, Dieuwke Luijten, Parham Sadeghipour, Julie Bayley, Scott C. Woller

**Author notes:** **Corresponding Author:** Samarth Mishra, BS^1^. **Senior Author:** Scott C. Woller, MD^9^.

## Abstract

**Background:** Physician communication with patients is a key aspect of excellent care. Scant evidence exists to inform best practice for physician communication in patients diagnosed with pulmonary embolism and deep vein thrombosis, collectively referred to as venous thromboembolism (VTE). The aim of this study was to summarize the existing literature on best practices for communication between healthcare providers and patients newly diagnosed with VTE.

**Methods:** We performed a scoping review of the extant literature on best practice for physician patient communication and the diagnosis and management of VTE. Manuscripts on communication between healthcare professionals and patients with acute vascular diseases, including VTE, were eligible. Two authors independently reviewed titles, and consensus determined article inclusion. The manuscripts were further categorized into two main categories: best practice in communication and unmet needs in communication. Data aggregation was achieved by a modified thematic synthesis.

**Results:** Among 345 initial publications, 22 manuscripts met inclusion criteria with 11 that addressed VTE, five pulmonary embolism, four deep vein thrombosis, one atrial fibrillation, and one acute coronary syndrome. Eleven manuscripts addressed communication of VTE diagnosis, while 12 focused on communication of VTE treatment. Eleven manuscripts identified unmet communication needs, and 14 addressed best practice. Our review shows that good communication surrounding the VTE diagnosis and treatment can enhance satisfaction while suboptimal communication can incur emotional, cognitive, behavioral, social, and health-systems adverse effects.

**Conclusion:** Scant literature guides best practices for communicating VTE diagnosis and treatment. Further research is necessary to establish practices for improving communication with VTE patients.

## Introduction and Background

Venous thromboembolism (VTE) includes pulmonary embolism (PE) and deep vein thrombosis (DVT). PE is the third leading cause of cardiovascular death, and the number one cause of preventable hospital-associated mortality [1, 2]. Increasingly, VTE is treated in an outpatient setting, which limits the time that healthcare providers can provide patient education and respond to patient’s questions. Physician-patient communication is a key influencer of patient perception of their current and future health state [3].

The physician-patient relationship is foundational for quality medical care. Effective physician-patient communication helps patients feel safe in stressful situations and instills confidence in the provider. However, pulmonary embolism and deep vein thrombosis are frequently diagnosed in the emergency department, which is a chaotic environment with brief encounters that may predispose to distress for many. Furthermore, delivering the diagnosis of VTE is a form of breaking bad news, a known inhibitor to patient comprehension and the capacity to internalize and retain information. For other conditions such as a cancer diagnosis, significant evidence exists that patient-physician communication affects provider trust and influences patient anxiety. [4] And in diagnosis of chronic diseases such as diabetes, high-quality communication between healthcare providers and patients is associated with better patient wellbeing and self-care [4]. In contrast, little is known about how healthcare provider communication is associated with patient recovery and wellbeing following a diagnosis of VTE. Key aspects of decision making in thrombosis medicine include the discussion surrounding the balanced risk versus benefit of invasive procedures, usually in the immediate term of diagnosis and duration of anticoagulation therapy later on in care.

Our study had two aims. First, to summarize the existing literature on best practices for communication between healthcare providers and patients newly diagnosed with pulmonary embolism or deep vein thrombosis, and second to identify knowledge gaps for opportunity in optimizing VTE communication.

## Methods

### Scoping literature review

A database search of PubMed, CINHAL, Web of Science, and Cochrane Library Databases for MeSH terms, title/abstract terms, and key categories for manuscripts that addressed physician-patient communication and VTE was conducted from inception of the database through June 21^st^, 2023 (Original Search, Figure 1). Due to the paucity of relevant articles identified, the search was expanded to include other vascular conditions in addition to VTE (Enhanced Search, Figure 1). The search criteria are represented in Table 1. The search protocol was adherent with the methodological steps advised according to the Preferred Reporting Items for Systematic reviews and Meta-Analyses (PRISMA) extension for Scoping Reviews [5].

**Figure 1.**
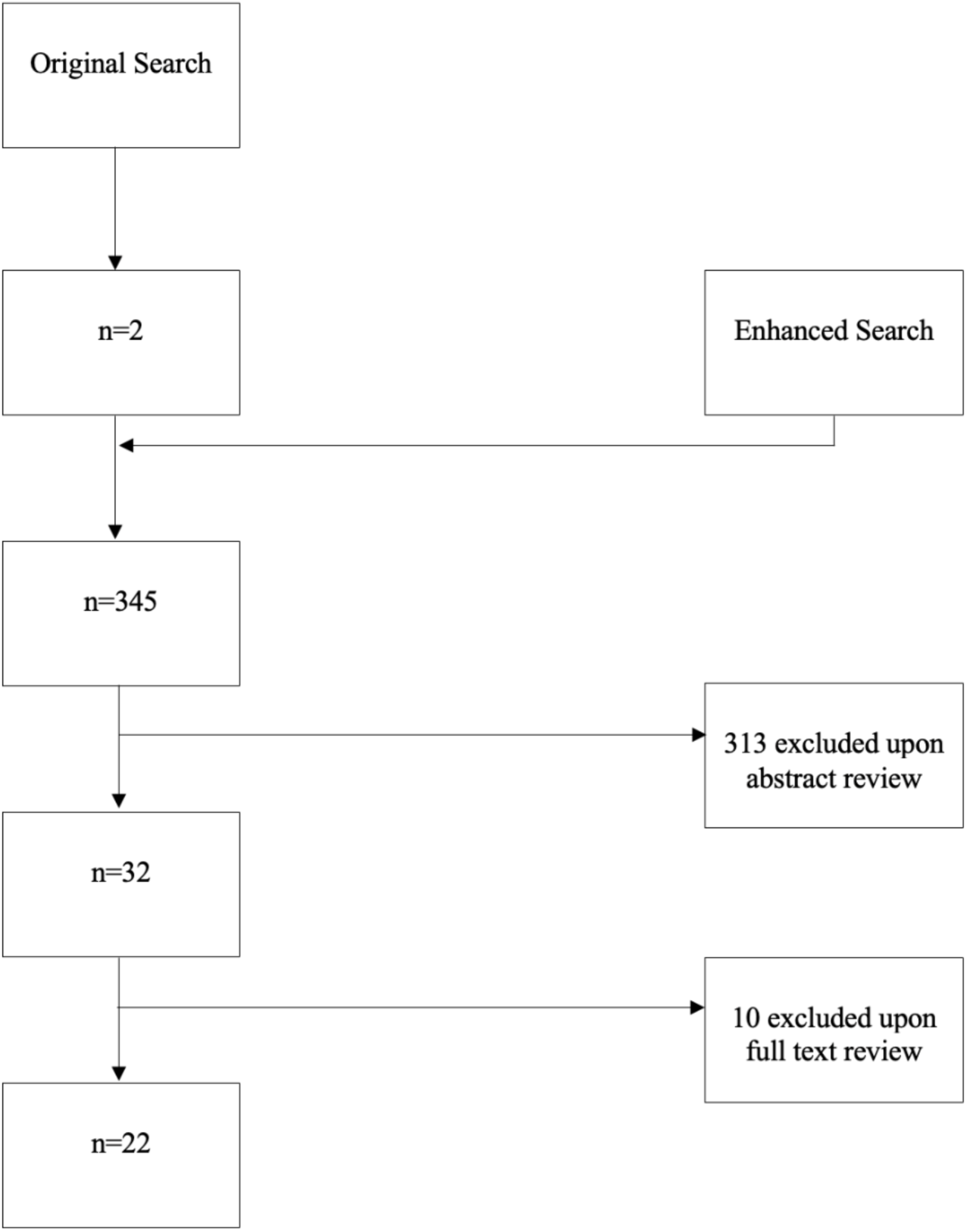
Flow Chart Scoping Review Search Methodology.

**Table 1.** Search Strategy with MeSH terms and Category Titles.

| **Table 1. Search Strategy with MeSH terms and Category Titles** |  |  |  |
| --- | --- | --- | --- |
| **Database** | **Search Criteria #1** | **Search Criteria #2** | **Search Criteria #3** |
| PubMed | "Venous Thrombosis"[Mesh] OR<br>"Pulmonary Embolism"[Mesh] OR | "Physician-Patient Relations"[Mesh] OR<br>"Nonverbal Communication"[Mesh] | "Vascular Diseases"[Mesh] |
|  | “DVT” OR “VTE” OR<br>“PE” OR “Deep Venous<br>Thromboses” OR “Deep<br>Venous Thrombosis” OR<br>"Venous<br>Thromboembolism"[Mesh] | OR "Interpersonal<br>Relations"[Mesh] OR<br>"Communication"[Mesh]<br>OR "Patient<br>Participation"[Mesh] OR<br>"Social<br>Participation"[Mesh] OR<br>"Psychology"[Mesh] |  |
| CINHAL | “MH Venous Thrombosis<br>+” | “MH Physician-patient<br>relations” OR (MH<br>"Psychology+") OR<br>"Engagement" | “MH Vascular<br>Diseases +” |
| Web of Science | “Venous Thrombosis” | “Physician-Patient<br>Relations” | “Vascular<br>Diseases” |
| Cochrane Library | "Venous<br>Thrombosis"[Mesh] OR<br>“Venous<br>Thromboembolism”<br>[Mesh] | “Physician-Patient<br>Relations” [Mesh] OR<br>“Psychology” [Mesh] | “Vascular<br>Diseases” [Mesh] |

### Eligibility criteria

Full-text manuscripts written in English, Spanish, German, Dutch, and French that addressed the topic of healthcare provider communication with patients experiencing acute VTE or vascular conditions were eligible for inclusion.

#### Study selection

Two authors (S.M., F.A.K) independently reviewed the titles and abstracts in this search. Disagreements were resolved initially by discussion and in the case of persistent disagreement a third author (S.C.W) adjudicated study inclusion by simple majority.

#### Data extraction and synthesis

Data aggregation was achieved by a modified thematic synthesis [6]. Two authors, (S.M., S.C.W) independently reviewed each manuscript and labeled each based on thematic focus.

Initial assessment of the articles led creation of four categories for descriptive purposes. These are presented in Table 2 with respective citations.

**Table 2.** Categorization of Included Manuscripts.

|  | **Primary Focus: VTE** | **Primary Focus: PE** | **Primary Focus: DVT** | **Primary Focus: AF** | **Primary Focus: ACS** |
| --- | --- | --- | --- | --- | --- |
| **Communication in VTE Diagnosis** | [8, 11, 13, 14, 25] | [12, 29, 30, 34] | [33] |  | [10] |
| **Communication in VTE Treatment** | [13, 16, 17, 22, 25, 37] | [12, 23, 34] | [15, 27] | [20] |  |
| **Unmet Needs in Communication [Table 4]** | [8, 11, 13, 14, 17, 22, 25, 37] | [12, 34] | [21] |  |  |
| **Best Practice in Communication [Table 5]** | [14, 16, 17, 31, 37] | [12, 23, 29, 30] | [15, 27, 33] | [20] | [10] |
| **VTE: venous thromboembolism, PE: pulmonary embolism, DVT: deep vein thrombosis, AF: atrial fibrillation, ACS: acute coronary syndrome** |  |  |  |  |  |

Manuscripts were categorized into areas of clinical focus that included venous thromboembolism (VTE), pulmonary embolism (PE), deep vein thrombosis (DVT), acute coronary syndrome (ACS), and atrial fibrillation. We further aggregated articles based on whether the manuscript addressed diagnosis or treatment. Lastly, we grouped articles based on which aspect of communication (identification of unmet needs, or provision of best practice) each addressed (Table 2).

## Results

The systematic search identified 345 manuscripts. A total of 32 manuscripts were identified for full-text analysis and 22 manuscripts met the inclusion criteria (Figure 1) [7–38].

Among the 22 included manuscripts, the primary focus of 11 was VTE (both PE and DVT), five PE, four DVT, one atrial fibrillation, and one acute coronary syndrome (ACS).

Eleven manuscripts addressed communication of the diagnosis, and 12 manuscripts addressed communication about treatment. Eleven manuscripts provided information surrounding the identification of unmet needs in communication and 14 provided guidance regarding best practice (see Table 2).

### Impact of physician-patient communication in VTE

Our review identified several areas of patient wellbeing influenced by physician communication in acute VTE and vascular diseases. de Wit et al. described that poor physician communication resulted in patient anxiety [8], and Etchegary et al. enumerated the “Zones of Relevance” that physician communication impacts as: emotional, cognitive, behavioral, social, and health services [11]. Furthermore, Genge et al. identified limitations of clinical trials in the VTE / vascular space and stated that the physical, emotional, and psychiatric impacts of VTE are beyond objective outcomes that are typically measured in clinical trials [13]. Existing evidence nevertheless suggests the importance of VTE diagnosis and treatment communication on patient psychological health. In an interview study, two of 72 PE patients experienced “ongoing and untreated psychological distress” that met the criteria for PTSD [34]. Patients who experienced ongoing psychological distress tended to recall poor communication at the time of diagnosis [34].

### Influence of health literacy on communication effectiveness in VTE

Several manuscripts point to patient health literacy and patient resource readability as central aspects of VTE communication. Fischer et al. proposed a role for questionnaires to “identify patients with low health literacy who may require additional support from the healthcare system…” [12]. San Norberto et al. identified that “venous thrombosis patient education materials produced by leading medical societies have readability scores that are above the recommended levels” [25]. The pivotal role of health literacy in patient satisfaction was described in a study of 2154 VTE patients receiving oral anticoagulation (OAC) treatment where “limited HL [health literacy] was associated with lower OAC treatment satisfaction…” [22].

Efforts in making VTE communication easily comprehendible and transmissible to the general population have shown success. Easy-to-understand communication to heighten public awareness of VTE through DVT public awareness campaigns have been found to increase DVT diagnosis rates [33].

### Timing and completeness of VTE communication

Evidence exists that physician communication at the time of the VTE diagnosis falls short of patient expectations. A study with telephone interviews among patients prescribed compression stockings (n=12) revealed that patients received little-to-no information regarding their compression stockings [21]. Likewise, a French study that included 103 VTE patients revealed that “more than 75% of patients reported that no physician warned them about risks of anticoagulation, long-term complications of venous thromboembolic disease or its prevention” [17].

### The importance of individualized approaches

A key aspect for effective communication and decision making in the acute diagnosis and management of VTE and vascular disease is the consideration of individual patient values and preferences. However, as evidenced in a systematic review of 48 studies conducted as part of a guideline review [20], values and preferences towards treatments vary, requiring more investment in understanding patient values in context. One study that focused on patient satisfaction with PE testing in the emergency department [30] found that patient satisfaction arises from addressing the patient’s primary concern, providing individualized care, utilizing imaging modalities for diagnosis (ex. CT scans), and from confidence from the treating physician [30]. A separate study of patient INR self-testing found that patient participation and engagement was more likely to be realized with personalized communication and direct physician-patient collaboration [16].

### The need for accurate disclosure and actively addressing difficult discussions

The communication of an acute VTE diagnosis can often involve numerous ethical variables, especially when decision-making may significantly impact a patient’s future quality of life. Patients identify that physician-patient collaboration in medical decision-making is an important satisfier, especially when stark differences in the possible outcomes exist [15]. A study centered around discussion of thrombolysis in the event of a submassive pulmonary embolism raised the importance of engaging patients in difficult conversations including conversations with ethical and legal implications; while difficult, this further engenders patient trust and satisfaction in final impressions regarding care [23]. Often difficult conversations include those that communicate medical error and unexpected, “incidental” findings. In a study of 971 patients that assessed patient satisfaction with VTE care, dissatisfaction was strongly associated with perceived mistakes in medical care, and the authors suggested that interventions aimed at reducing, acknowledging, and communicating errors might improve patient satisfaction in care [37]. Similarly, in a study assessing communication of incidental findings for another indication, Sonis et al. stated “communication of clinically significant incidental findings identified in the ED to patients and their primary care physicians is essential” [29].

### Specialized, but not role-restricted communication

Evidence exists that best practice for communication in the domain of acute VTE is a “team sport.” A Turkish study that included nurse-led DVT training on inpatient protective self-care practices found that nurse education improved patient comprehension [27]. Facilitating specialists in thrombosis endorsed that “information provided by vascular medicine physicians was clearer and more complete” when compared to a non-specialist physician [17].

### Active awareness of physician communication style and medicalization of information

Finally, best practice includes that the physician be mindful of overt (patient health literacy or language fluency) aspects of communication and indirect factors of communication (e.g., cultural sensitivities). Hernandez-Nino et al. identified that aspects of a healthcare encounter may inadvertently incite fear over reassurance [14] when a physician’s word choice (use of medical jargon), behavior, balance of fear versus reassurance, and the amount of information provided [14] come up short. Edelman et al. evaluated the impact of native language on the care of patients in the acute-care setting. He stated, “…native language concordance acts as a protective factor for patient-clinician interpersonal care in the acute setting, regardless of native language or English proficiency” [10]. Literature has shown that it is attainable to provide information and care to patients that they are able to comprehend rather easily. Taking Takara et al. as an example, an informative manual was made on VTE for the lay population with 97.5% of readers deeming the manual as easily understandable and 90.0% of readers finding the reading not tiresome [31].

Manuscripts that discussed unmet needs in communication are referenced in Table 4, and manuscripts that discussed best communication practices are referenced in Table 5.

**Table 4.** Manuscripts detailing Unmet Needs in Communication.

| **Article Title** | **Author (s)** | **Country** | **Year** | **Study/**<br>**Article Type** | **Summative Statement** |
| --- | --- | --- | --- | --- | --- |
| “What information patients require on graduated compression stockings.” [21] | May, V., et al. | England | 2006 | Qualitative – patient interviews/ surveys | n=12 study indicating that patients who were using compression stockings had little or no information about their stockings. |
| “Patient Satisfaction With Venous Thromboembolism Treatment” [37] | Webb, D., et al. | U.S.A. | 2019 | Qualitative – patient interviews/ surveys | Perceived mistakes during care for VTE was strongly associated with VTE care dissatisfaction. |
| “Roles of the general practitioner and the vascular medicine physician for patient education concerning venous thromboembolism: The patient’s perspective.” [17] | Le Collen, L., et al. | France | 2019 | Qualitative – patient interviews/ surveys | Calls for standardization of VTE patient education and communication of essential educational messages pertaining to VTE. |
| “Communication at diagnosis of venous thromboembolism: Lasting impact of verbal and nonverbal provider communication on patients.” [14] | Hernandez-Nino, J., et al. | U.S.A. | 2022 | Qualitative – patient interviews/ surveys | Indicates that nonverbal communication and utilization of medical jargon can influence a patient’s response and can, in some cases, have a negative impact on patient-provider communication. |
| “The psychological impact of pulmonary embolism: a mixed-methods study” [34] | Tran, A., et al. | Canada | 2021 | Mixed methods | 2 of 72 patients were found to meet the diagnosis of PTSD due to ongoing psychological distress following a PE diagnosis. |
| “Health literacy in patients with pulmonary embolism: development and validation of the HeLP (Health Literacy in Pulmonary Embolism) – Questionnaire.” [12] | Fischer, S., et al. | Germany | 2023 | Mixed methods | Proposed the development of an assessment tool to evaluate patient health literacy and which patients may require additional healthcare support. |
| “Psychosocial aspects of venous thromboembolic disease: an exploratory study.” [11] | Etchegary, H., et al. | Canada | 2008 | Exploratory | VTE diagnosis communication can impact “Zones of Relevance” which includes emotional, cognitive, behavioral, social, health services. Patient knowledge deficits in the VTE setting raises questions to current knowledge provisions and consent protocols. |
| “Do physicians contribute to psychological distress after venous thrombosis?” [8] | De Wit, K. | Canada | 2022 | Commentary | Mismatch between physician goals and patient needs in the VTE setting is contributing to increased prevalence of patient anxiety amongst patients with VTE. Mental recovery of patients can be improved with thought, empathy, and good communication skills. |
| “Evaluation of patients’ experience and related qualitative | Genge, L., et al. | Canada | 2022 | Scoping Review | Clinical trials studying VTE often are unable to measure the physical, emotional, and psychiatric components that come alongside a VTE diagnosis. |
| outcomes in venous thromboembolism: a scoping review” [13] |  |  |  |  |  |
| “Health Literacy and Treatment Satisfaction Among Patients with Venous Thromboembolism” [22] | Mefford, M. T., et al. | U.S.A. | 2022 | Retrospective Cohort | Patients with limited health literacy were found to have lower oral anticoagulant treatment satisfaction. |
| Readability of patient educational materials in venous thrombosis: analysis of the 2021 ESVS guidelines and comparison with other medical societies information.” [25] | San Norberto, E.M., et al. | Spain | 2022 | Cross-sectional | Readability of VTE patient education materials by leading societies were found to have readability scores above the recommended levels. |

**Table 5.** Manuscripts detailing Best Communication Practices.

| Article Title | Author (s) | Country | Year | Study Type | Summative Statement |
| --- | --- | --- | --- | --- | --- |
| “Technology-assisted self-testing and management of oral anticoagulation therapy: a qualitative patient-focused study.” [16] | Kuljis, J., et al. | U.K. | 2016 | Qualitative – patient interviews/ surveys | Patient engagement and involvement is likely to be realized more when patients receive timely, individualized, and face-to-face care. |
| “Patient Satisfaction With Venous Thromboembolism Treatment” [37] | Webb, D., et al. | U.S.A. | 2019 | Qualitative – patient interviews/surveys | Mentions importance in disclosing any reasons for treatment mistakes, diagnostic mistakes, and/or delayed treatment to preserve a healthy patient-physician relationship |
| “Roles of the general practitioner and the vascular medicine physician for patient education concerning venous thromboembolism: The patient’s perspective.” [17] | Le Collen, L., et al. | France | 2019 | Qualitative – patient interviews/surveys | With respect to VTE diagnosis communication, patients reported to have better communication from vascular, specialized physicians when compared to general physicians. Calls for a standardized approach for VTE communication. |
| “Patient values and preferences in pulmonary embolism testing in the emergency department” [30] | Swarup, V., et al. | Canada | 2021 | Qualitative – patient interviews/surveys | Patients being worked up for PE in the ED were found to prefer care that was individualized, centered around their primary concern, and care that involved utilization of medical imaging. |
| “Communication at diagnosis of venous thromboembolism: Lasting impact of verbal and nonverbal provider communication on patients.” [14] | Hernandez-Nino, J., et al. | U.S.A. | 2022 | Qualitative – patient interviews/surveys | Utilization of simple terminology and single components is likely to improve VTE communication in the acute care setting. Nonverbal communication and expressions should be an additional area of focus. |
| “Incorporating patients’ preferences into medical decisions.” [15] | Kassirer, J.P. | U.S.A. | 1994 | Commentary | Patients should play an active role in medical decision making – especially when decisions may have differences in possible outcomes. |
| “Communication of imaging recommendations to ED patients: Pulmonary embolus CT.” [29] | Sonis, J. D., et al. | U.S.A. | 2016 | Commentary | Communication of incidental findings is essential to pave a pathway for patients to trust their physicians and follow-up/attain further care. |
| “Patient values and preferences in decision making for antithrombotic therapy: a systematic review” [20] | MacLean, S., et al. | Canada | 2012 | Systematic Review | Patient values and preferences with respect to treatment vary due to prior experiences or health outcomes. Patient preferences should be considered prior to initiating any treatments. |
| “Effect of a public awareness campaign on the incidence of symptomatic objectively confirmed deep vein thrombosis: a controlled study.” [33] | Tomkowski, W. Z., et al. | U.S.A. | 2012 | Observational – Case-control study | Raising public awareness for DVT resulted in increased diagnosis. When awareness campaign was halted, diagnosis rates dropped to baseline. |
| “Controversy and consent: achieving patient autonomy in thrombolysis for acute submassive pulmonary embolism.” [23] | Pywell, S. R., et al. | U.K. | 2015 | Case Report | Refers to the presentation of a patient with PE and discussion of the difficulties associated with attaining consent. Ultimately, when patient and physician have disagreement over treatment options, crucial to involve the patient further for the final decision. |
| “Nurse-led patient training improves deep vein thrombosis | Serpici, A.; Gursoy, A. | Turkey | 2018 | Quasi-experimental | Vital to incorporate all members of the medical team together when treating a patient. Study found that when patients were educated and trained by nurses, they had |
| knowledge and self-care practices.” [27] |  |  |  |  | more knowledge and took more responsibility for their own care. |
| “Development and validation of an informative manual on venous thromboembolism for the lay population” [31] | Takara, N.C., et al. | Brazil | 2020 | Methodological Study | Development of an informative manual on VTE was found to be understood by 97.5% of readers with another 90.0% stating the reading was not tiresome. |
| “Impact of Native Language, English Proficiency, and Language Concordance on Interpersonal Care During Evaluation of Acute Coronary Syndrome.” [10] | Edelman, D. S., et al. | U.S.A. | 2022 | Prospective Cohort | Language concordance serves as a protective factor for the patient-clinician relationship in the acute-care setting. |
| “Health literacy in patients with pulmonary embolism: development and validation of the HeLP (Health Literacy in Pulmonary Embolism) – Questionnaire.” [12] | Fischer, S., et al. | Germany | 2023 | Mixed methods | Vital to consider patient health literacy when providing care to patients with VTE. |

## Discussion

As VTE communication improves, patient experience and satisfaction will, too. However, the literature that informs best practice in communication surrounding acute VTE diagnosis and treatment is limited. To the best of our knowledge, this is the first comprehensive review summarizing existing literature to inform best practice in acute VTE diagnosis and treatment. A method of modified thematic synthesis was employed to summarize manuscripts that met inclusion criteria. Thematic synthesis is a method of data extraction that permits aggregations of themes, and has been described in studies that address questions of patient perspective and experience [39]. Our method differed from that formerly described by Thomas and Harden [6] as we adopted a synthesis of the principal theme of the manuscript, as opposed to a “line by line” coding that was out of scope for our review.

These findings inform future clinical and research implications. Our results inform that clinical practice should focus on enhancing the clinician-patient relationship, especially when delivering a VTE diagnosis. These findings were not exclusive to communication between patients and physicians but extended to all members of a patient’s care team, including nurses. Our review shows that good communication surrounding the VTE diagnosis and treatment can enhance satisfaction and engagement while suboptimal communication can incur emotional, cognitive, behavioral, social, and health-systems adverse effects. Patient health literacy and understanding patient preference are cornerstones of effective communication. This study serves as the foundation for determining how to operationalize these conditions in emergency departments and ambulatory care to embed the benefits of expert communication in routine care delivery models. Specific direction including a “physician best-practice toolkit in VTE communication” are yet to be established. Further research and literature surrounding beneficial communication practices when delivering a VTE diagnosis needs to be further explored.

Our results are inherently limited by the paucity of literature that informs best practice in communication surrounding VTE diagnosis and treatment. Our results were limited to those obtained using MeSH terms and category titles across four databases. Despite the expansion of the search criteria to other vascular conditions, few (22) articles met the inclusion criteria for this study. Though our data may be limited in providing specific communication practices for a VTE diagnosis, our study proves useful in identifying unmet needs in VTE diagnosis and treatment.

## Conclusion

We conducted a scoping review of the extant literature regarding communication by the physician to the patient of the acute diagnosis and treatment of VTE. The results of this review indicate that opportunity exists to establish a best practice for physician-patient communication in the setting of acute VTE diagnosis and treatment. Limited literature exists to inform specific communication recommendations for delivering a VTE diagnosis. Understanding the existing guidance and knowledge gaps can serve as a roadmap to create a physician VTE communication toolkit that might provide all providers an evidence-based resource to optimize communication in the acute diagnosis and treatment of VTE.

## Data Availability

The data used to support the findings of this study are included within the article.

